# DETERMINING DECISION THRESHOLDS FOR PHYSICIANS AND PARENTS FOR INSTITUTING A DO-NOT-ATTEMPT-RESUSCITATION ORDER FOR PEDIATRIC IN-HOSPITAL CARDIAC ARREST PATIENTS – A CROSS-SECTIONAL STUDY OF PARENTS AND PHYSICIANS IN THE UNITED STATES

**DOI:** 10.64898/2026.02.17.26346477

**Authors:** Minaz Mawani, Ye Shen, Jessica Knight, Bryan McNally, Mark Ebell

**Affiliations:** Epidemiology and Biostatistics, University of Georgia College of Public Health, Athens, Georgia; Larner College of Medicine, University of Vermont Department of Medicine, Burlington, Vermont; Department of Family Medicine, Michigan State University, East Lansing, Michigan; Emory University School of Medicine, Atlanta, Georgia

**Keywords:** Resuscitation orders, Critical illness, Child, Counseling

## Abstract

**Background and Objectives:** Decision-making about resuscitating a critically ill child is complex yet common. We aimed to study the survival thresholds at which physicians, compared to parents, decide to treat or withhold resuscitating a child. Moreover, we aimed to compare physicians’ survival estimates with those from a nationwide registry.

**Methods:** We conducted a cross-sectional survey-based study in the United States. Clinical vignettes based on hypothetical survival probabilities were used to study and compare the decision thresholds for parents and physicians. Vignettes developed using the Get-With-The-Guidelines-Resuscitation registry were used to explore physicians’ decision thresholds and compare their survival estimates with those from the data. Thresholds were determined using mixed-effect logistic regression models.

**Results:** We had decisions for 501 and 257 vignettes from 167 parents and 43 physicians, respectively. The decision threshold for survival to discharge was 5.3% (95% CI: 3.7 to 7.0) for physicians and 1.2% (95% CI: −0.8 to 3.0) for parents. Whereas the decision threshold for survival to discharge with PCPC 1 or 2 was 3.5% (95% CI: 1.1 to 7.1) for physicians and 0.6% (95% CI: −1.2 to 1.8) for parents. About 58% of the physicians overestimated the likelihood of survival.

**Conclusions:** The study found that the decision threshold for the physicians was higher than that for the parents (5.3% vs. 1.2%). This illustrates that parents still want to attempt resuscitation at a survival probability where physicians would recommend withholding resuscitation. These findings have implications for clinical practice and counseling the parents of critically ill hospitalized children.

## Introduction

The decision to terminate resuscitative efforts in pediatric cardiopulmonary arrest is complex, multifactorial, and not uncommon.^1^ As many as 68% of deaths in Dutch children between 0 and 1 year of age were preceded by an end-of-life decision.^2^ In addition, most deaths in pediatric intensive care units (PICU) follow either withdrawal or limitation of life-sustaining treatments, and the number of these patients has increased over time.^3,4,5^

Common reasons to choose withdrawal of life-sustaining therapy include prolonged invasive therapy, fear of the child developing a brain injury, and financial and emotional burdens.^6,7^ End-of-life decision making requires a physician’s ability to correctly estimate survival. Ebell and colleagues’ research determined that physician’s ability to correctly predict survival in adults following cardiac arrest was not better than chance alone (Area under the receiver operating characteristic curve = 0.47).^8^ However, it is not known whether these findings would be different in children. Another study found that the threshold to suggest withdrawal of life-sustaining therapy based on chances of survival or projected quality of life differs between intensivists.^9^ Moreover, these decisions are also affected by non-medical factors such as parental absence at the bedside, physician’s years of clinical experience, their moral and ethical perspectives, their definition of a good quality of life for a child, and personal experiences such as a child reminding a physician of their family member.^1^

The survival thresholds at which a physician decides to treat or withhold resuscitating a child have never been estimated.^9,10^ Studies that claim to guide determinations of futility among critically ill patients are based on insufficient data to provide any statistical confidence for clinical decision-making, which increases the degree of subjectivity.^11^ There are currently no prediction models or risk scores to identify children for whom any resuscitation would be futile as opposed to adults where several scores have been developed to predict survival outcomes.^12–24^

Therefore, we aimed to determine the threshold level of survival to discharge and neurological outcome below which a Physician would not want to resuscitate and how this level differs from a parent’s threshold. We also compared physicians’ self-estimated survival with the model-based survival calculated using a clinical dataset and compared their decisions before and after the provision of model-based probabilities.

## Methods

### Study Design and Population

This was a cross-sectional survey-based study; physicians and parents from across the United States were recruited using convenience sampling. We included physicians who have cared for hospitalized children in the United States since completing residency training and parents living in the United States. We used multiple strategies to recruit participants described below.

#### Parents

Parents were contacted and invited to participate through the ResearchMatch registry. ResearchMatch is a large Web-based, national health volunteer recruitment registry that was created by several academic institutions and supported by the U.S. National Institutes of Health as part of the Clinical Translational Science Award (CTSA) program.^25^ We also invited potential participants through social media (Facebook, NextDoor), and snowball sampling through friends and family.

#### Physicians

For recruiting physicians, we reached out to potential participants through our contacts at university hospitals (Children’s Hospital of Philadelphia, Children’s Hospital of Atlanta, Emory University etc.), networks of physicians (North American Primary Care Research Group, Pediatric Resuscitation Quality Collaborative investigators, Pediatric Acute Lung Injury and Sepsis Investigators network, Pediatric Section of Society of Critical Care Medicine etc.), and social media applications. We shared our recruitment material (flyer, Qualtrics survey link, and recruitment message) around Facebook and Twitter using relevant hashtags (#pedsICU, #PedsCICU, #neoTwitter, #societyofcriticalcaremedicine, #CriticalCare, #SCCMSoMe, #PhDVoice, #phdchat, #AcademicTwitter, @AmerAcadPeds, and @PhDVoice).

### Survey Questionnaire

We developed secure web-based surveys for parents and physicians using Qualtrics, provided through the University of Georgia (UGA). We did not collect any identifying information, including IP addresses; all records were given a unique study ID, multiple entries by participants were blocked, and the data were password protected. The study was reviewed and deemed exempt by the Institutional Review Board (IRB) at the UGA.

#### Pilot Testing

We conducted a pilot test with physicians and parents to evaluate the clarity of language and time required for completion. Based on the responses, we modified the survey to make it easier to understand and complete. For instance, we initially had three different treatment options to select from: Full code, DNAR (Do-Not-Attempt-Resuscitation), or if they would like more information to decide. Most participants selected the third option, which would have made our study results inconclusive. Therefore, we modified it to only 2 options. We also added simplified descriptions of medical terminologies such as ‘Cardiac Arrest’, ‘Cardiopulmonary Arrest’, ‘DNAR’, and ‘Neurological survival’. We also modified the survey to improve language sensitivity. We did not use the responses from pilot testing in our main study data analysis.

#### Patient and Public Involvement

Patients or the public were not involved in the design, or conduct, or reporting, or dissemination plans of our research except for the pilot testing of questionnaire.

#### Main Study Data

An individual could choose to participate in the study after reviewing the consent letter. Participants were asked questions about demographics and their decisions for a child who has undergone cardiac arrest. Our survey questionnaire had two different parts. A set of identical vignettes to be completed by parents as well as physicians (Questionnaire I) and a set of vignettes to be completed by physicians alone (Questionnaire II). The details are given below.

#### Identical Vignettes for Physicians and Parents (Questionnaire I)

We developed six different vignettes using hypothetical survival and neurologically intact survival probabilities. Each participant (parents and physicians) was presented with three vignettes of a child admitted with a critical illness in an intensive care unit (Appendix, part II). Participants were then asked to choose one of two options: to keep the child as full code or to consider a do-not-attempt-resuscitation order. We simplified the language of the vignettes for parents to an 8th-grade level. A summary for the survival and neurologically intact survival probabilities, and participants’ decisions is given in Table. I, Appendix. Participants were also asked about their definition of good neurological outcome – Pediatric Cerebral Performance Category (PCPC = 1, PCPC = 1 or 2 or PCPC = 1, 2, or 3)

#### Vignettes for Physicians Alone (Questionnaire II)

We developed an additional 9 vignettes with survival probabilities calculated using a logistic regression model based on data from the “Get With The Guidelines Resuscitation” (GWTG-R) Registry (Appendix, part I). Inpatients who received a documented episode of cardiopulmonary resuscitation (CPR) were included. Patients who received CPR in the operating room or procedure suite were excluded. Each physician was given 3 randomly selected vignettes to evaluate (Appendix part II). We asked physicians to estimate the survival probability (0 to 100%) if this child experiences an in-hospital cardiac arrest episode. In addition, they were asked whether they would recommend full aggressive management or suggest that parents consider a do-not-resuscitate option based on their estimated survival probability. We later provided the model-based survival probability for those vignettes and asked the physicians to decide again based on this newly provided survival probability. Self-estimated and model-based probabilities, as well as decisions based on each probability, were compared.

### Data Analysis

All analyses were performed using R (version 2023.06.0; The R Foundation, Vienna, Austria). We summarized the characteristics of parents and physicians and their decisions using mean with standard deviation, median with interquartile range, and numbers with percentages. We compared the characteristics of participants across the categories of decision using a chi-square test or an independent sample t-test. When a range of survival probabilities were provided by the participants, we used the midpoint of the range for instance 5% for 0 – 10%.

We applied the threshold model of clinical decision-making to in-hospital cardiac arrest vignettes to determine a single decision threshold for DNAR vs full code. Below the DNAR decision threshold survival is deemed unlikely enough that a DNAR order is suggested, while above the threshold survival is likely enough that the patient should be full code (Figure 1). The survival probability at which 50% of the physicians and parents suggest DNAR status and the other half choose to provide full code is the ‘DNAR decision threshold’.^26^ ^27–31^

**Figure 1:**
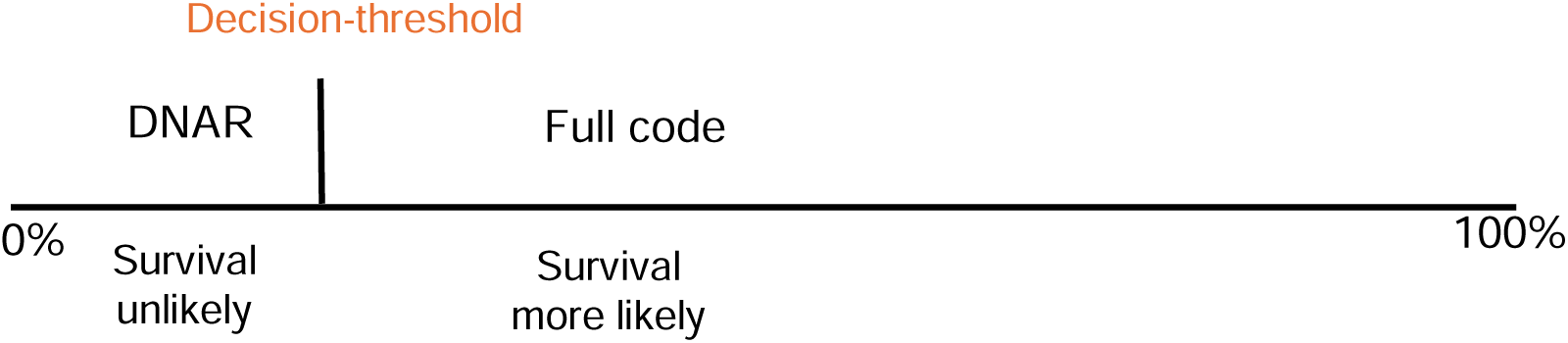
Illustration of the threshold model of decision-making, showing a single decision threshold for DNAR vs. Full code.

#### Comparing Physician and Parent Decision Thresholds for DNAR Orders (Questionnaire I)

Considering that each physician and parent evaluated several vignettes, we used mixed effects logistic regression model and introduced a random intercept term into the equation to adjust for repeated observations. The DNAR decision thresholds were estimated using the function glmer() from the package lme4 in R. Confidence intervals were calculated using the variance-covariance matrix associated with the logistic regression model. We also adjusted for other variables such as age, gender, clinical experience, education level, etc., to compare the DNAR decision thresholds for subgroups of populations (Appendix, Part I: details on calculation).

#### Comparing Physician Estimates of Survival with Those of a Multivariate Model (Questionnaire II)

The physician estimate of survival was compared with the model-based survival probability using a calibration plot. Decision Thresholds were calculated using a mixed effect logistic regression model. Reclassification tables were also used to present physician decisions before and after provision of the model-based survival probability.

## Results

A total of 171 parents and 43 physicians responded to the invitation. Of these 167 parents and 43 physicians provided usable data. Most of the parents had a bachelor’s or a master’s degree. Most physicians (58%) were trained in pediatric critical care, and most (74%) had more than 10 years of clinical experience. The mean age for parents and physicians was similar. Compared to parents, fewer physicians were women (75% vs. 61%, p = 0.001). The Southern US was the most common geographic region (63% of physicians and 44% of parents, p = 0.01). Parents were less likely to choose a DNAR decision compared to physicians (44% vs. 64%, p = <0.0001). There were no other differences in the demographic characteristics of the two groups (Table 1). Overall, parents provided their decisions for a total of 501 vignettes and physicians for 257 vignettes.

**Table 1:** Demographic characteristics of participating parents (n = 167) and physicians (n = 43).

| **Characteristics** | **n (%)** |
| --- | --- |
| ***Parents*** |  |
| Mean age (SD), range (years) | 48.8 (12.4), 24 to 81 |
| Female | 124 (74.2) * |
| Region |  |
| Northeast | 22 (13.2) |
| Midwest | 47 (28.1) |
| South | 73 (43.7) |
| West | 25 (14.9) |
| Level of education |  |
| High school, college, or associate's degree | 35 (20.9) |
| Bachelor's degree | 41 (24.5) |
| Master's degree | 71 (42.5) |
| Doctoral degree | 20 (11.9) |
| Their definition of a good neurological outcome |  |
| PCPC 1 | 4 (2.4) |
| PCPC 1 or 2 | 21 (12.7) |
| PCPC 1, 2, or 3 | 140 (84.8) |
| ***Physicians*** |  |
| Mean age (SD), range (years) | 47.4 (10.5), 30 to 70 |
| Female | 26 (60.5%) |
| Mean years of practice (SD), range | 20.5 (11.3), 5 to 44 |
| Region |  |
| Northeast | 8 (18.6) |
| Midwest | 2 (4.6) |
| South | 27 (62.7) |
| West | 6 (13.9) |
| Clinical Specialty |  |
| Neonatologist | 5 (11.6) |
| Pediatric critical care physician | 26 (60.4) |
| Pediatric cardiologist | 2 (4.6) |
| Pediatric emergency physician | 1 (2.3) |
| Family medicine | 4 (9.3) |
| Other specialty** | 5 (11.6) |
| Their definition of good neurological outcome |  |
| PCPC 1 | 0 (0) |
| PCPC 1 or 2 | 9 (20.9) |
| PCPC 1, 2, or 3 | 34 (79.1) |

SD: Standard deviation, PCPC: Pediatric cerebral performance category, \*2 parents described their gender as non-binary, \*\* 1 each of nephrology, rheumatology, pulmonary critical care, child abuse pediatrics, infectious diseases

### Comparing Physician and Parent Decision Thresholds for DNAR Orders (Questionnaire I)

The percentage of physicians recommending a DNAR ranged from 40% to 85%. Whereas, for parents, the percentage deciding for DNAR ranged from 28% to 56% (Table 1, Appendix). The decision threshold for survival to discharge was 5.3% (95% CI: 3.7 to 7.0) for physicians and 1.2% (95% CI: −0.8 to 3.0) for parents. The decision thresholds for neurologically intact survival to discharge (PCPC = 1 or 2) were 3.5% (95% CI: 1.1 to 7.1) for physicians and 0.6% (95% CI: −1.1 to 1.8) for parents. (Figures 2a and 2b). Thresholds for survival to discharge were lower (e.g. less likely to write a DNAR order) for physicians who were older than 50 years, had more than 10 years of experience, or were men. There was no significant difference in decision thresholds by region or specialty. There were no statistically significant differences in the decision thresholds for parents across the categories of age, gender, region, education (Table 2).

**Figure 2a:**
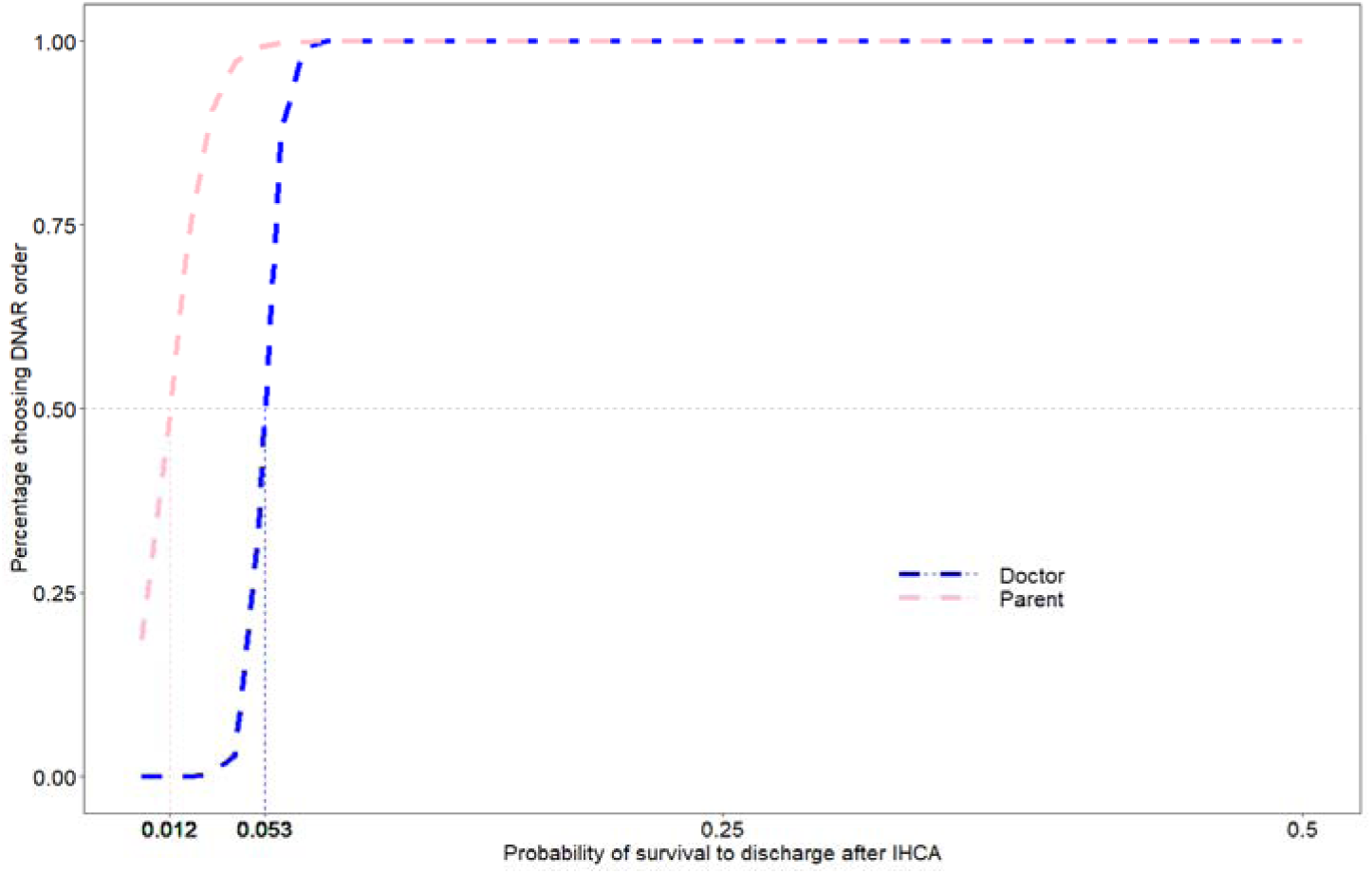
Plots of decision thresholds for physicians and parents for overall survival to discharge after IHCA. The threshold is the point where the curve crosses the horizontal 50% bar.

**Figure 2b:**
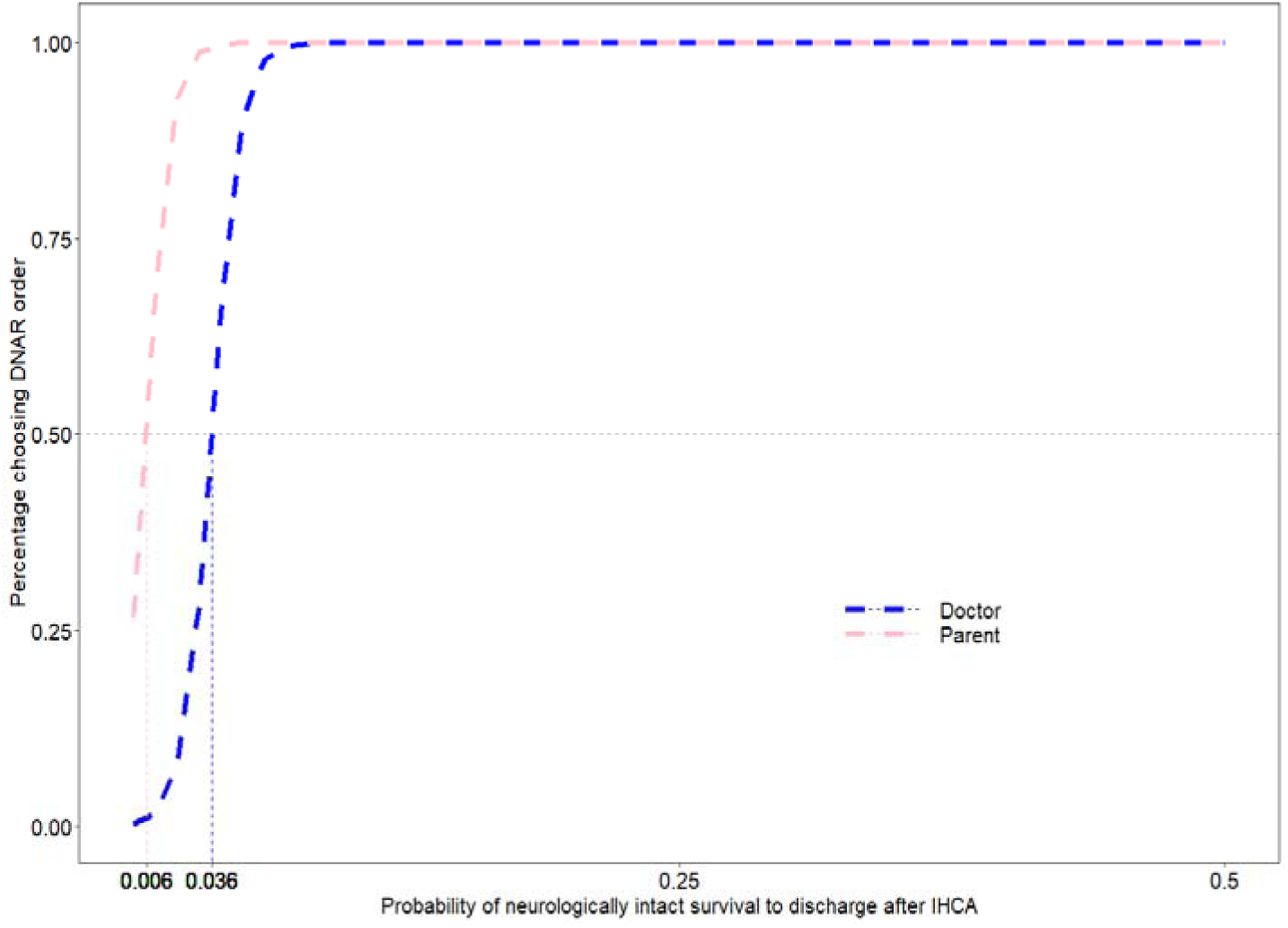
Plots of decision thresholds for physicians and parents for neurologically intact survival to discharge after IHCA. The threshold is the point where the curve crosses the horizontal 50% bar.

**Table 2:** Estimation of DNAR decision thresholds using hypothetical vignettes and survival probabilities, by subgroups with 95% CIs for parents and physicians (Questionnaire I).

|  | Any survival to discharge |  | Survival with PCPC = 1 or 2 |  |
| --- | --- | --- | --- | --- |
| Subgroup | DNAR decision threshold (95%CI) | P-values | DNAR decision threshold (95%CI) | P-values |
| **Parents** |  |  |  |  |
| All parents | 1.2 (-0.8 to 3.0) |  | 0.6 (-1.1 to 1.8) |  |
| Gender |  | 0.69 |  | 0.67 |
| Female | 1.2 (-1.1 to 3.3) |  | 0.6 (-1.2 to 2.0) |  |
| Male | 0.5 (-3.4 to 3.9) |  | 0.0 (-2.9 to 2.4) |  |
| Age |  | 0.41 |  | 0.41 |
| Younger <=50 | 0.6 (-2.1 to 2.9) |  | 0.1 (-2.2 to 1.8) |  |
| Older > 50 | 2.0 (-0.9 to 4.7) |  | 1.1 (-1.2 to 3.0) |  |
| Region |  | 0.45 |  | 0.44 |
| South | 0.5 (-2.5 to 3.1) |  | 0.1 (-2.4 to 1.9) |  |
| Others | 1.8 (-0.8 to 4.2) |  | 1.0 (-1.1 to 2.7) |  |
| Education |  | 0.07 |  | 0.07 |
| Up to bachelors | 0.5 (-3.6 to 2.1) |  | 0.6 (-3.1 to 1.2) |  |
| Master's and above | 2.6 (0.1 to 5.0) |  | 1.6 (-0.4 to 3.3) |  |
| **Physicians** |  |  |  |  |
| All physicians | 5.3 (3.7 to 7.0) |  | 3.5 (1.1 to 7.1) |  |
| Gender |  | <0.001 |  | 0.37 |
| Female | 5.6 (3.9 to 7.7) |  | 4.2 ( -6.8 to 13.5) |  |
| Male | 4.4 (1.7 to 7.4) |  | 2.7 (-9.4 to 12.9) |  |
| Age |  | <0.001 |  | 0.44 |
| Younger <=50 | 5.7 (3.8 to 8.2) |  | 4.0 (1.1 to 8.7) |  |
| Older > 50 | 4.6 (2.2 to 7.2) |  | 2.7 (-1.6 to 7.1) |  |
| Region |  | 0.92 |  | 0.84 |
| South | 5.3 (3.5 to 7.3) |  | 3.6 (0.7 to 8.2) |  |
| Others | 5.2 (2.3 to 8.2) |  | 3.3 (-1.1 to 8.3) |  |
| Years of experience |  | 0.02 |  | 0.08 |
| <= 10 years | 8.5 (6.5 to 11.7) |  | 5.9 (3.8 to 9.4) |  |
| >10 year | 4.5 (3.0 to 6.6) |  | 2.9 (1.5 to 4.2) |  |
| Specialty |  | 0.61 |  | 0.60 |
| Pediatric critical care | 5.6 (3.7 to 7.8) |  | 3.6 (2.0 to 6.4) |  |
| Others | 4.8 (2.4 to 7.5) |  | 3.1 (1.2 to 5.1) |  |

DNAR: Do-not-attempt resuscitation, PCPC: Pediatric cerebral performance category

### Comparing Physician Estimates of Survival with Those of a Multivariate Model (Questionnaire II)

Only 8 (6.2%) of the physicians’ decisions were estimated within +/- 20% of the model-based probability of survival; 74 (58%) overestimated the probability of survival to discharge and 46 (36%) underestimated it (Figure 3). There were no differences in physician’s characteristics compared across the categories of overestimation (Appendix Table IV).

**Figure 3:**
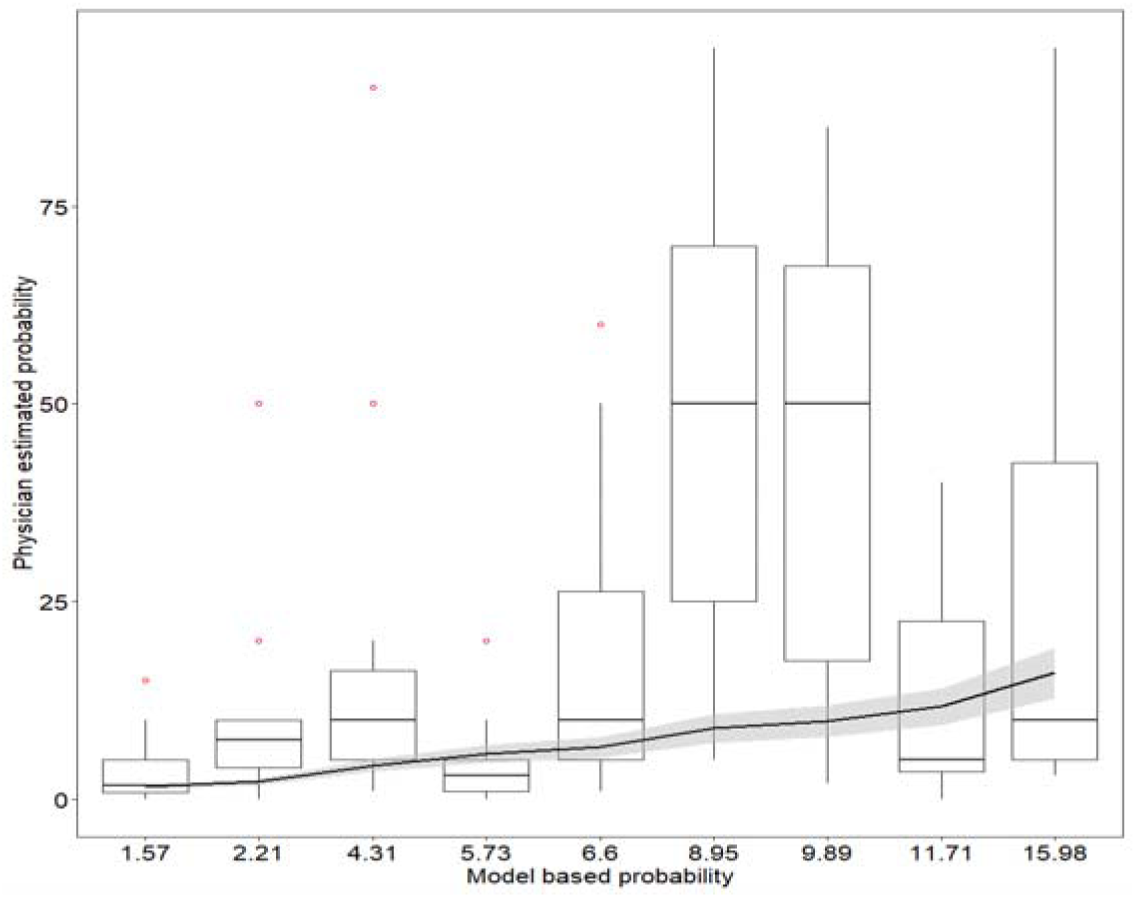
Physician-estimated survival probability compared to model-based probability for each clinical vignette. Boxes represent the median, interquartile range, and overall range of survival estimates for each of the 9 distinct clinical vignettes. The thick black line represents perfect agreement, and the grey shaded region represents +/-20% of the model-based probability (Questionnaire II).

Table 3 summarizes the data on reclassification following the provision of model-based survival probabilities. Among 128 vignettes for which a second decision was also made, 108 (84.3%) had the same decision before and after the provision of the model-based probability despite the large amount of overestimation. Only in 7 (5.4%) cases, the decision changed from full code to DNAR. In 13 (10.1%) cases, the decision changed from DNAR to full code.

**Table 3:** Reclassification table showing the relationship between decisions made based on physician-estimated probability of survival to discharge following IHCA (initial decision) and the updated decision once presented with the multivariate model-based probability of survival (Questionnaire II).

|  | Decision based on model-based probability |  | Total |
| --- | --- | --- | --- |
| Decision based on physician-estimated probability | Full code | DNAR |  |
| Full code | 63 | 7 | 70 |
| DNAR | 13 | 45 | 58 |
| Total | 76 | 52 | 128 |

\*DNAR: Do-not-attempt-resuscitation

The DNAR decision threshold based on the physician-estimated survival probability was 13% (95% CI: 6.6 to 18.9) and the DNAR decision threshold for physicians after being given the model-based probability was 5.3% (95% CI: 2.7 to 7.1). The latter is the same as the thresholds identified using hypothetical survival probabilities.

## Discussion

This is the first study to explore DNAR decision thresholds for parents and physicians for pediatric in-hospital cardiac arrests. Using a set of hypothetical probability-based vignettes, the decision thresholds for overall survival and neurologically intact survival (PCPC = 1 or 2) for physicians were estimated to be 5.3% and 3.6%. For parents, the thresholds were considerably lower at 1.2% and 0.57%. Parents would still want to attempt resuscitation at a survival probability where physicians would choose a DNAR. This is an important finding from a clinical practice perspective and supports involving parents in the decision making of a critically ill child. Physicians and parents are known to have different understanding of the outcomes.^32^ Parents need prognostic information to develop realistic expectations for their child, and participating in shared decision-making as surrogates for their child.^33^ While there is a general understanding to involve parents in the decision-making for their hospitalized children, persistent gaps are observed in patient-provider interaction.^34^ This has implications for counseling and educating parents in a better way and explaining to them the prognosis and potential harms of resuscitation procedures.

Our study did not explore the causes of discrepancy between a physician’s and parent’s thresholds. A study identified that most surrogates were reluctant to accept physician’s predictions of medical futility believing that it may be unreliable, wanting to see for themselves the futility of the situation and a belief in miracles or divine action. Involving the ethicist may help understand the nature of the conflict and better communicate with the parents. If a religious belief is the basis of conflict, it would be useful to involve chaplain or a representative of the surrogate’s religion, besides a good communication from physicians.^35^

In our study the decision thresholds were lower for physicians that were older, had more than 10 years of experience or were men. Previous research suggests quite the opposite, as physicians gain more experience, they are better able to recognize futility and more likely to make DNAR decisions.^1,36–38^ Our findings may be attributed to random error because of the small sample size and a study with a larger sample size may help test the hypotheses.

Our study also found that physicians most often overestimated the probability of survival. Similar findings were shared by another study, which also found that the physicians were unable to discriminate between patients with and without a better chance of survival following resuscitation.^39^ The probability of survival estimated by physicians might influence parent’s choice about resuscitating their child and therefore it has implications for clinical practice. The model-based cases were deliberately selected to reflect patients with a low probability of survival, to enhance our ability to detect a threshold which may have contributed to the overestimation of survival by physicians.

Our findings support the need for a tool to help with the estimation of survival and thereby standardize decision making in critical care. We have developed such a tool based on state-wide registry data that can potentially help with the process (unpublished data). The DNAR decision threshold for survival to discharge estimated using hypothetical vignettes as well as model-based vignettes, was 5.3%, which shows that our findings are consistent and have good face validity.

### Strengths

To the best of our knowledge, this is the first study exploring the decision thresholds for parents and physicians for hospitalized children giving insights about DNAR decision making and the difference between physicians and parents in their approach. We collected data from across the US and also used a national registry which makes our study results generalizable. We used very low probability of survival in our vignettes which was helpful in identifying the very low survival group which poses a dilemma for physicians to decide a DNAR or full code.

### Limitations

Calculating decision thresholds is a useful and objective way to look at clinical decision-making, however, the process is more complex and multifactorial than a binary logistic regression model could gauge. It may involve other factors that we did not have access to, including physiological parameters, details regarding prognosis for specific conditions, and other factors. We also cannot exclude the possibility of social desirability bias. Our sample size for physicians was small and may need another larger study to support these findings.

## Conclusion

Using model-based and hypothetical probability of survival, we estimated decision thresholds for physicians and parents. Half of physicians would consider opting for a DNAR below a survival of 5.3%, and half of parents would consider a DNAR below a survival of 1.2%. These findings have implications for clinical practice, especially counseling the parents of critically ill hospitalized children.

## Data Availability

All data produced in the present study are available upon reasonable request to the authors

## Acknowledgements

We thank our colleagues and friends for completing the pilot survey and sharing their insights that helped us modify the survey. We thank all the participants for completing the survey and helping us understand this very important topic. We thank physicians from the pediRES-Q network and different hospitals in the US for taking the time from their busy schedules to take an interest in the topic, sharing insights from their clinical experiences, and completing the survey. We especially thank Dr. Maryam Naim for reviewing the protocol and providing critical feedback. We thank Yewen Chen for his help with programming.

## Conflict of interest disclosures (includes financial disclosures)

None

## Author Contributions

Dr. Minaz Mawani conceptualized and designed the study, acquired and analyzed the data, drafted the initial manuscript, and critically reviewed and revised the manuscript.

Dr. Mark Ebell conceptualized and designed the study, helped with the analysis and interpretation of the data, and critically reviewed and revised the manuscript.

Dr. Ye Shen helped with the analysis and interpretation of data and critically reviewed and revised the manuscript.

Dr. Jessica Knight critically reviewed and revised the study protocol and manuscript Dr. Bryan McNally critically reviewed and revised the study protocol and manuscript

All authors approved the final manuscript as submitted and agree to be accountable for all aspects of the work.

## APPENDIX

### I. Model for calculating survival to discharge for a child (1 yr to <12 yrs)

Ln(p/1-p) = 0.45 - 0.98 (Illness Category - Medical Cardiac) - 0.93 (Illness Category – Medical Non-Cardiac) - 0.24 (Illness Category – Surgical Cardiac) + 0.55 (Acyanotic Cardiac Malformation) - 0.99 (Hepatic Insufficiency) – 0.53 (Hypotension/Hypoperfusion) – 0.05 (Metabolic or Electrolyte Abnormality) – 0.89 (Metastatic or Hematologic Malignancy) – 0.55 (Renal Insufficiency) – 0.6 (Septicemia) + 0.3 (PCPC 1 or 2)

The ‘DNAR decision thresholds’ were determined by adopting the method described in a previously published work.^27^ This method is based on logistic regression analysis of the DNAR decisions for parents and physicians. The following logistic regression equation was used.

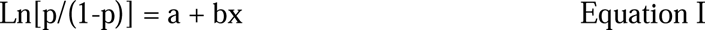

p is the probability of recommending a do-not-resuscitate status. x is the hypothetical probability from the vignettes. The threshold probability is defined as the probability X such that the corresponding probability of withholding or treating is equal to 0.5. Whereas x is the hypothetical survival probability. Solving equation I for threshold, the final equation I was x = - (a/b) as ln (0.5/0.5) = 0.

### II. Sample of Clinical Vignettes

**Part (i) Example of vignettes using hypothetical survival probabilities. We used identical vignettes for parents and physicians for this section (Questionnaire I).**

**Parent’s Questionnaire**

The purpose of this section is to learn about decision-making for critically ill children from a parent’s perspective. For each of the scenarios given below, please select your decision from the given options. Please select only one response for each scenario.

You have been selected to review scenarios 1 - 3.

**<u>Scenario 1</u>:** Imagine that you are the parent of a previously healthy **5-year-old child** who is now **critically ill** due to a serious lung infection. He has been admitted to the **intensive care unit** and is on a mechanical ventilator (a machine that breathes for him). He is not doing well, and his doctors say that they would expect only **3 out of 100** children who are this ill to survive. However, there will only be a **1 in 100** chance that your child will have the same level of intelligence and function after his illness even if he does survive. The doctors ask whether you would like them to try to **revive** him using CPR (chest compressions and shocks) if his **heart stops**. Your **decision** is:

- You ask them to try to **revive** him if his heart stops
- You ask that a **do-not-resuscitation** order be placed to avoid unnecessary suffering **<u>Scenario 2</u>:** *Same as scenario 1* but his doctors say that they would expect only **7 children out of 100** who are this ill to survive. Furthermore, there is also only a **5 in 100** chance that your child will have the same level of intelligence and function after his illness even if he does survive. Your **decision** is:
- You ask them to try to **revive** him if his heart stops
- You ask that a **do-not-resuscitation** order be placed to avoid unnecessary suffering
- **<u>Scenario 3</u>:** *Same as scenario 1* but doctors say that they would expect only **5 out of**

**1000** children who are this ill to survive. Furthermore, there is only a **3 in 1000** chance that your child will have the same level of intelligence and function after his illness even if he does survive. Your **decision** is:

- You ask them to try to **revive** him if his heart stops
- You ask that a **do-not-resuscitation** order be placed to avoid unnecessary suffering

**Physician’s Questionnaire**

For each of the scenarios mentioned below consider yourself as a physician caring for the patient in the scenario. The patient’s parents ask you for your recommendation regarding CPR for their child. Please select one of the following options as your recommendation for each scenario.

You have been selected to review scenarios 1 - 3.

**<u>Scenario 1:</u>** Imagine that you have a previously healthy **5-year-old child** under your care who is now **critically ill** due to a serious lung infection. He has been admitted to the **intensive care unit** and is on a mechanical ventilator. Based on a validated prediction model and your evaluation, he is not doing well and has only a **3%** chance of survival and a **1%** chance of a good neurological outcome if he survives. What will be your **recommendation** to the child’s parents regarding a do-not-attempt-resuscitation order compared to a full code?

- You recommend that the child be on **full code** and receive **CPR**
- You recommend that the parents consider a **do-not-attempt-resuscitation** order for this child

**<u>Scenario 2</u>:** *Same as scenario 1* but has a **7%** chance of survival and a **5%** chance of a good neurological outcome if he survives. What will be your **recommendation** as the child’s physician to the parents?

- You recommend that the child be on **full code** and receive **CPR**
- You recommend that the parents consider a **do-not-attempt-resuscitation** order for this child

**<u>Scenario 3</u>:** *Same as scenario 1* but has a **0.5%** chance of survival and a **0.3%** chance of a good neurological outcome if he survives. What will be your **recommendation** as the child’s physician to the parents?

- You recommend that the child be on **full code** and receive **CPR**
- You recommend that the parents consider a **do-not-attempt-resuscitation** order for this child

Part (ii) Example of vignettes from physician’s questionnaire using model-based probabilities (Questionnaire II).

For each clinical scenario below, enter the probability of survival and choose your preferred recommendation.

**<u>Scenario 1</u>:** Imagine that you have a **5-year-old child** under your care who was admitted for the management of **congenital heart disease**. He has been admitted to the **intensive care unit** and has **hepatic insufficiency**, **hypotension**, **hematologic malignancy**, **renal insufficiency**, and **septicemia**. The child was admitted with a **good PCPC** (pediatric cerebral performance category) score (1 or 2). If this child undergoes **cardiac arrest**, what do you think are the **chances** that the child will **survive to discharge** from the hospital? (Enter your estimate of survival probability from 0 to 100%)

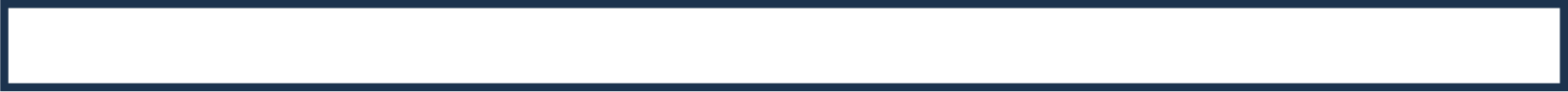

For the above scenario, what will be your **recommendation** as a child’s physician to the parents.

- You recommend that the child be on **full code** and receive **CPR**
- You recommend that the parents consider a **do-not-attempt-resuscitation** order for this child

If you are told that the child in the above scenario has a **2.21%** chance of survival to discharge, what will be your **recommendation** as a child’s physician to the parents?

- You recommend that the child be on **full code** and receive **CPR**
- You recommend that the parents consider a **do-not-attempt-resuscitation** order for this child

**Table I:** Survival, neurological outcome, and comparison of decisions by parents and physicians for each vignette based on hypothetical probabilities (Questionnaire I).

| Case Scenario | Survival to discharge | Neurologically intact survival | DNAR / Total (%) |  | P-value |
| --- | --- | --- | --- | --- | --- |
|  |  |  | Physician's decision | Parents decision |  |
| 1 | 0.03 | 0.01 | 14/20 (70) | 34/73 (46.6) | 0.06 |
| 2 | 0.07 | 0.05 | 8/20 (40) | 26/73 (35.6) | 0.71 |
| 3 | 0.005 | 0.003 | 17/20 (85) | 41/73 (56.2) | 0.02 |
| 4 | 0.05 | 0.03 | 12/23 (52.2) | 26/94 (27.7) | 0.02 |
| 5 | 0.02 | 0.01 | 13/23 (56.5) | 45/94 (47.9) | 0.45 |
| 6 | 0.01 | 0.006 | 18/23 (78.3) | 48/94 (51.1) | 0.02 |

DNAR: Do-not-attempt-resuscitation

**Table II:**
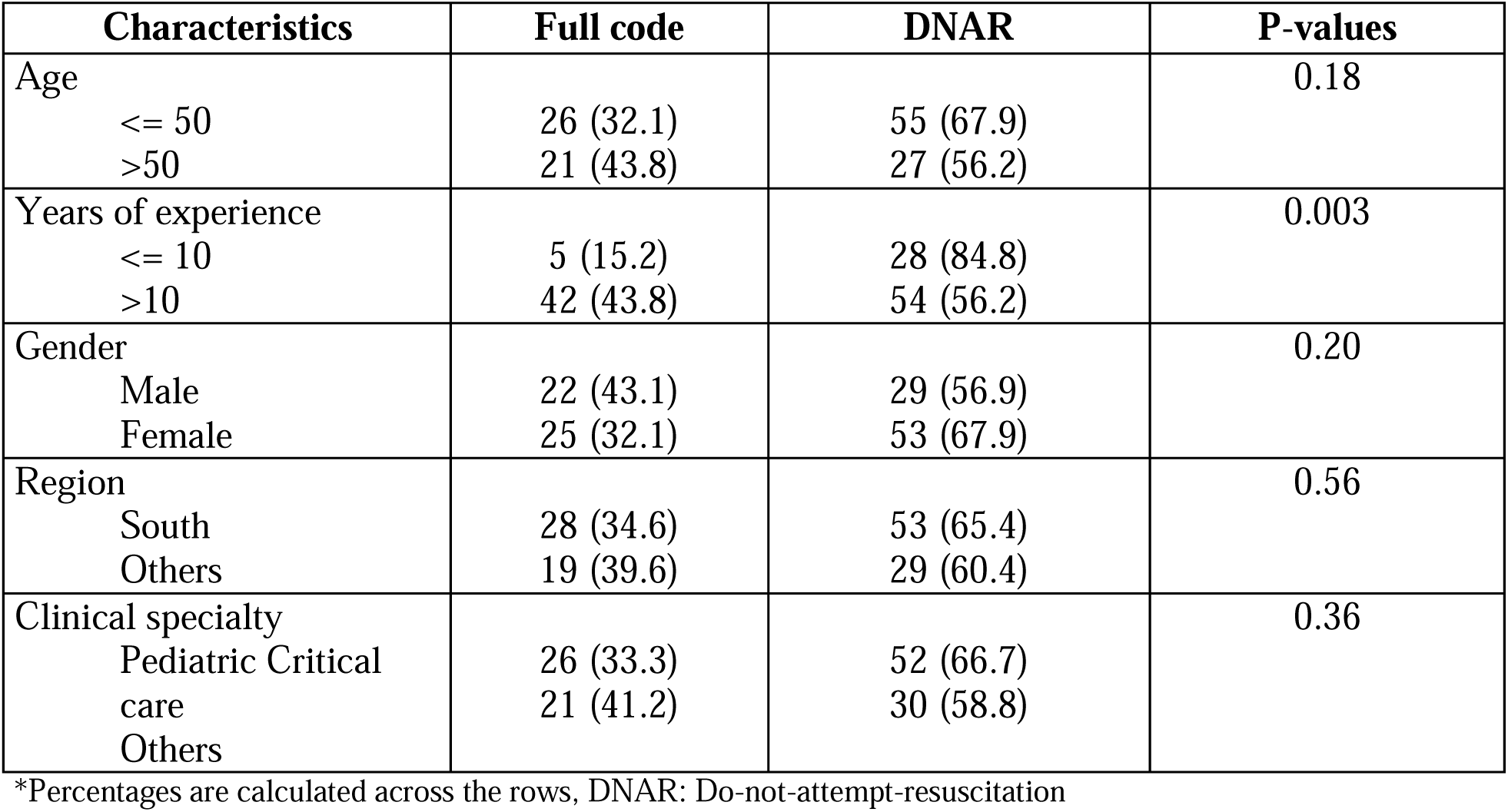
Comparison of decisions of physicians across all vignettes using identical vignettes for physicians and parents consisting of hypothetical survival probability (Questionnaire I).

| Characteristics | Full code | DNAR | P-values |
| --- | --- | --- | --- |
| Age |  |  | 0.18 |
| <= 50 | 26 (32.1) | 55 (67.9) |  |
| >50 | 21 (43.8) | 27 (56.2) |  |
| Years of experience |  |  | 0.003 |
| <= 10 | 5 (15.2) | 28 (84.8) |  |
| >10 | 42 (43.8) | 54 (56.2) |  |
| Gender |  |  | 0.20 |
| Male | 22 (43.1) | 29 (56.9) |  |
| Female | 25 (32.1) | 53 (67.9) |  |
| Region |  |  | 0.56 |
| South | 28 (34.6) | 53 (65.4) |  |
| Others | 19 (39.6) | 29 (60.4) |  |
| Clinical specialty |  |  | 0.36 |
| Pediatric Critical care | 26 (33.3) | 52 (66.7) |  |
| Others | 21 (41.2) | 30 (58.8) |  |

\*Percentages are calculated across the rows, DNAR: Do-not-attempt-resuscitation

**Table III:** Comparison of decisions of parents across all vignettes using identical vignettes for physicians and parents consisting of hypothetical survival probability (Questionnaire I).

| Characteristics | Full code | DNAR | P-values |
| --- | --- | --- | --- |
| Age |  |  | 0.19 |
| ≤ 50 | 167 (58.6) | 118 (41.4) |  |
| >50 | 114 (52.8) | 102 (47.2) |  |
| Gender |  |  | 0.64 |
| Male | 72 (58.5) | 51 (41.5) |  |
| Female | 209 (56.2) | 163 (43.8) |  |
| Region |  |  | 0.19 |
| South | 130 (59.4) | 89 (40.6) |  |
| Others | 151 (53.5) | 131 (46.5) |  |
| Level of Education |  |  | 0.006 |
| Up to bachelors | 143 (62.7) | 85 (37.3) |  |
| Master's and PhD | 138 (50.5) | 135 (49.5) |  |

\*Percentages are calculated across the rows, DNAR: Do-not-attempt-resuscitation

**Table IV:**
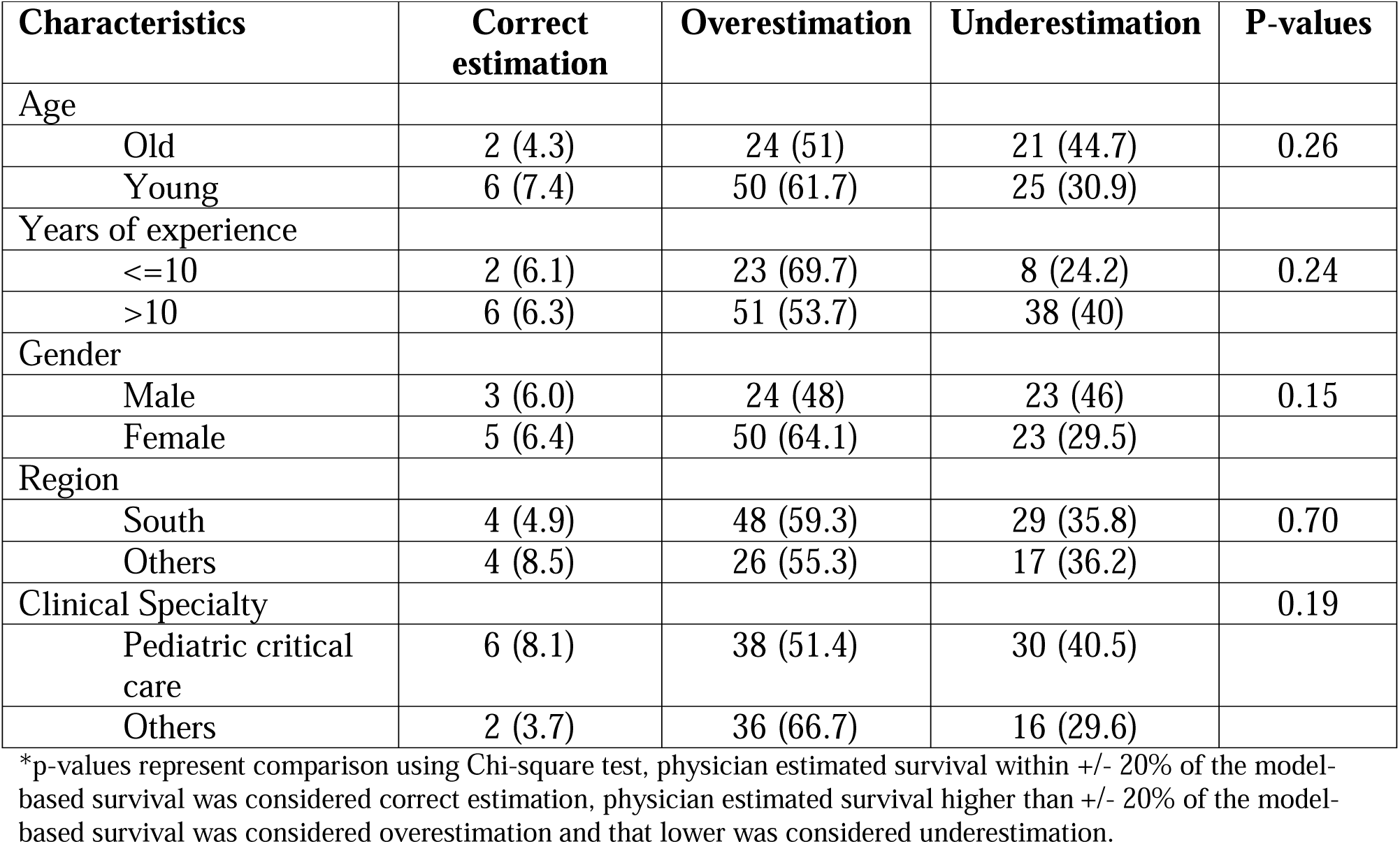
Comparing overestimation across physician characteristics using physician-estimated survival probability and model-based survival probability (Questionnaire II).

| Characteristics | Correct estimation | Overestimation | Underestimation | P-values |
| --- | --- | --- | --- | --- |
| Age |  |  |  |  |
| Old | 2 (4.3) | 24 (51) | 21 (44.7) | 0.26 |
| Young | 6 (7.4) | 50 (61.7) | 25 (30.9) |  |
| Years of experience |  |  |  |  |
| ≤10 | 2 (6.1) | 23 (69.7) | 8 (24.2) | 0.24 |
| >10 | 6 (6.3) | 51 (53.7) | 38 (40) |  |
| Gender |  |  |  |  |
| Male | 3 (6.0) | 24 (48) | 23 (46) | 0.15 |
| Female | 5 (6.4) | 50 (64.1) | 23 (29.5) |  |
| Region |  |  |  |  |
| South | 4 (4.9) | 48 (59.3) | 29 (35.8) | 0.70 |
| Others | 4 (8.5) | 26 (55.3) | 17 (36.2) |  |
| Clinical Specialty |  |  |  | 0.19 |
| Pediatric critical care | 6 (8.1) | 38 (51.4) | 30 (40.5) |  |
| Others | 2 (3.7) | 36 (66.7) | 16 (29.6) |  |

\*p-values represent comparison using Chi-square test, physician estimated survival within +/- 20% of the model-based survival was considered correct estimation, physician estimated survival higher than +/- 20% of the model-based survival was considered overestimation and that lower was considered underestimation.

### III. Consent letters for Physicians and Parents

**UNIVERSITY OF GEORGIA CONSENT LETTER**

Determining decision thresholds to institute a Do-Not-Attempt Resuscitation (DNAR) status for a pediatric in-hospital cardiac arrest patient – a cross-sectional survey.

**Researcher’s Statement**

My name is Minaz Mawani, and I am a doctoral candidate in the Department of Epidemiology and Biostatistics at the University of Georgia under the supervision of Dr. Mark Ebell, Professor of Epidemiology. I am inviting you to take part in a research study. The information in this form will help you decide if you want to be in the study.

**Purpose of the Study**

We are conducting a survey to understand the decision-making process for physicians regarding resuscitating a child who is critically ill and may experience cardiac arrest in a hospital setting.

**Study Procedures**

If you agree to participate, you will be asked to

- Complete a short survey. The survey will inquire about your demographic information. You will also be asked to review different scenarios and decide as a physician, whether or not you would opt for a cardiopulmonary resuscitation for the child in the scenario. We will not collect any identifiable information. The questionnaire will take up to 10 minutes to complete.
- The scenarios will describe a patient’s condition, such as their diagnosis, procedure, clinical condition, and predicted survival, which can help the decision-making process.

**Risks and discomforts**

The foreseeable discomforts include psychological risks (e.g., feelings of stress/discomfort, sadness guilt, or anxiety related to any past experience of loss) for which you can decline to participate or are free to leave during the study.

**Benefits**

There are no direct benefits to you for completing the survey. However, your participation in the study will help us understand the decision-making process for children undergoing cardiac arrest. With the increasing number of Do-not-attempt-resuscitation decisions in this population, it is important to understand the difference between the decisions of parents vs. doctors in these scenarios. Having no previously published data on this topic, our study will help contribute to scientific knowledge. It may also help improve the decision-making process, and thereby allocation of resources in critical care settings and in turn benefit society.

**Privacy/Confidentiality**

Your privacy and confidentiality will be protected in the following ways.

-Personal identifiers will not be collected, and all records will be given codes to differentiate them from other records.

-The data will be password protected and no one else except for the study team members will have access to that. The records may be reviewed by departments at the University of Georgia responsible for regulatory and research oversight.

-Any publications resulting from this project will show aggregate results and will not identify you as a participant.

-This research involves the transmission of data over the Internet and there are some limits to data security and confidentiality with the use of online data collection methods, however, we will keep your responses confidential and use them for only the purpose of this research.

-Furthermore, the records will not be shared with other researchers or used for projects beyond this one.

Taking part is voluntary

Your involvement in the study is voluntary. You can refuse to participate or withdraw your consent at any time without giving any reason and without penalty or loss of benefits to which you are otherwise entitled.

If you have questions

If you are interested in participating or have questions about this research, please feel free to contact me at or Dr. Mark Ebell at. If you have any questions or concerns regarding your rights as a research participant in this study, you may contact the Institutional Review Board (IRB) Chairperson at 706.542.3199 or.

By completing this survey, you are consenting to participate in this study.

Sincerely,

Minaz Mawani

**UNIVERSITY OF GEORGIA CONSENT LETTER**

**Understanding the decision-making process for critically ill children that undergo cardiac arrest in the hospital - a cross-sectional survey.**

**Researcher’s Statement**

My name is Minaz Mawani, and I am a doctoral candidate in the Department of Epidemiology and Biostatistics at the University of Georgia under the supervision of Dr. Mark Ebell, Professor of Epidemiology. I am inviting you to take part in a research study. The information in this form will help you decide if you want to be in the study.

**Purpose of the Study**

We are conducting a survey to understand the decision-making process for parents regarding resuscitating a child who is critically ill and may experience cardiac arrest in a hospital setting.

Cardiac arrest is when a child’s heart suddenly and unexpectedly stops pumping. If this happens, blood stops flowing to the brain and other vital organs. The child becomes unconscious and stops breathing.

’Cardiopulmonary resuscitation’ (CPR) or ‘resuscitation’ is an emergency intervention performed by compressing the chest and providing artificial breaths which keep blood and oxygen flowing to the brain until the heart starts to beat again. Sometimes it also involves giving electric shocks to the heart also known as ‘defibrillation’. Usually, the health care team will try to revive the child by providing CPR unless the family has chosen a do-not-attempt-resuscitation for the child.

A do-not-attempt-resuscitation (DNAR) is a medical order which implies that the child should not receive CPR in the event that their heart stops beating. It is ethically acceptable to not resuscitate a child when it is unlikely to be successful in restoring heart function or when the risks are more than the benefits, to avoid unnecessary suffering.

Sometimes with advanced illness, CPR may be able to save the life but because of lack of circulation to the brain and complicated disease process, the child may have brain damage and need to stay on life support devices and medications and may not have the same level of function and quality of life when they go home.

**Study Procedures**

If you agree to participate, you will be asked to complete a short survey. The survey will ask about your basic information such as age, gender, education, etc. We will not collect any information that identifies you. You will also be asked to review different scenarios and decide whether as a parent you would decide to resuscitate (provide CPR to) the child in the scenario or choose not to. The questionnaire will take up to 10 minutes to complete.

**Risks and discomforts**

The foreseeable risk includes emotional discomfort (e.g., feelings of stress, sadness guilt, or anxiety related to any experience of loss) for which you can decline to participate or are free to leave during the study.

**Benefits**

There are no direct benefits to you for completing the survey. However, your participation in the study will help us understand the decision-making process for children undergoing cardiac arrest. It will also help us understand the difference between the decisions of parents vs. doctors in these scenarios.

**Privacy/Confidentiality**

Your privacy and confidentiality will be protected in the following ways.

-Personal identifiers (any information that identifies you as an individual) will not be collected, and all records will be given codes to differentiate them from other records.

-The data will be password protected and no one else except for the study team members will have access to that. The records may be reviewed by departments at the University of Georgia responsible for regulatory and research oversight.

-Any publications resulting from this project will show aggregate results and will not identify you as a participant.

-This research involves the transmission of data over the Internet and there are some limits to data security and confidentiality with the use of online data collection methods, however, we will keep your responses confidential and use them for only the purpose of this research.

-Furthermore, the records will not be shared with other researchers or used for projects beyond this one.

**Taking part is voluntary**

Your involvement in the study is voluntary. You can refuse to participate or withdraw your consent at any time without giving any reason and without penalty or loss of benefits to which you are otherwise entitled.

**If you have questions**

If you are interested in participating or have questions about this research, please feel free to contact me at or Dr. Mark Ebell at. If you have any questions or concerns regarding your rights as a research participant in this study, you may contact the Institutional Review Board (IRB) Chairperson at 706.542.3199 or.

**By completing this survey, you are consenting to participate in this study.**

Sincerely,

Minaz Mawani

